# Long-term impact of pre-natal exposure to the Ukraine famine of 1932-1933 on adult type 2 diabetes mellitus

**DOI:** 10.1101/2023.12.02.23299317

**Authors:** L.H. Lumey, Chihua Li, Mykola Khalangot, Nataliia Levchuk, Oleh Wolowyna

## Abstract

**Importance:** The long-term impacts of early-life famine exposure on Type 2 Diabetes Mellitus (T2DM) have been widely documented across countries, but it remains less clear what is the critical time window and if there is a dose-response between famine intensity and risk of T2DM.

**Objective:** To establish the relation between prenatal famine exposure and adult Type 2 diabetes mellitus (T2DM).

**Design:** A national cross-sectional study.

**Setting:** The man-made Ukrainian Holodomor famine of 1932-1933.

**Participants:** A total number of 128,225 T2DM cases diagnosed at age 40 or over from the national diabetes register 2000-2008 in Ukraine. The population at risk includes 10,186,016 Soviet Ukraine births (excepting one oblast/province) between 1930-1938 classified by month and year and oblast of birth.

**Exposure:** Births born in January-June 1934 from oblasts that experienced extreme, severe, or significant famine in 1932-1933. Famine intensity was measured based on the excess mortality during the famine.

**Main Outcomes and Measures:** T2DM diagnosis was based on WHO (1999) criteria.

**Results:** We observed in univariate analysis a 1.8-fold increase in T2DM (OR 1.80; 95% CI 1.74-1.85) among individuals born in the first half-year of 1934 in regions with extreme, severe, or significant famine. We observed no increase among individuals born in regions with no famine. In multivariate analysis across regions and adjusting for season of birth we observed a larger than 2-fold increase (OR 2.21; 95% CI 2.00-2.45). There was a dose-response by famine intensity, with ORs increasing from 1.94 to 2.39 across regions. The pattern was similar in men and women.

**Conclusions and Relevance:** Births in the first half-year of 1934 were conceived at the height of the Ukraine famine in 1933. This relation for T2DM outcomes points to early gestation as a critical time window relating maternal nutrition in pregnancy to offspring health in later life. Further studies of biological mechanisms should focus on this time window for which changes in DNA methylation and later body size have also been observed.

**Key Points:** *Question:* What is the critical time window for early-life famine exposure on Type 2 Diabetes Mellitus (T2DM)? Is there any dose-response relationship between early-life famine exposure and T2DM?

*Findings:* We found an increased risk of T2DM (Odds Ratio 2.21; 95% CI 2.00-2.45) among individuals born during the first half-year of 1934. A clear dose-response relationship was observed using excess mortality as a measure of famine intensity.

*Meaning:* Early gestation is a critical time window relating maternal undernutrition in pregnancy to offspring health in later life.

## INTRODUCTION

Russia’s deliberate war strategy in Ukraine has disrupted agriculture and food storage and distribution systems to hinder food supplies to the population, with consequences extending beyond Ukraine.^1,2^ The war has created an immediate humanitarian crisis of catastrophic proportions with uncertain long-term health effects.^1^ In 1932-1933, Ukrainian food supplies were also deliberately obstructed by Soviet interventions, leading to about 4 million excess deaths in the short-term.^3^ This famine in Ukraine is called Holodomor (death by starvation) to underline these events. We report on the long-term impact of the Holodomor on Type 2 Diabetes Mellitus (T2DM) cases diagnosed seven decades after prenatal famine exposure.

To study long-term health effects of early famine, large and well-defined birth cohorts are needed with significant variations in intensity of famine exposure.^4,5^ In addition, health outcomes need to be documented using standardized measures. The Ukraine setting provides an unusual opportunity to investigate this question because the famine was concentrated in a six-month period in early 1933, it showed extreme variations in intensity across oblasts (provinces), and adult outcomes were assembled in a national diabetes registry without consideration of place and date of birth. For T2DM in later life, a regional study of the Ukraine famine together with local studies of the Dutch Famine of 1944-45, the Chinese Famine of 1959-61 and three famines in 20th century Austria all suggest a relation with prenatal nutrition.^6-11^ We previously reported a 1.5-fold increase in T2DM in Ukraine among men and women born in four oblasts with extreme famine and a 1.3-fold increase in three oblasts with severe famine.^9^ As potential limitations we noted that the population at risk for T2DM was defined by survivors included in the 2001 Ukraine census and that the study covered only nine out of 24 oblasts.

Three improvements in this study allow for a more definitive estimate of the relation between early famine timing and T2DM. First, we identify the study population at birth rather than from survivors included in the census. Second, we expand the study to cover births in all but one of 24 Ukraine oblasts instead of being limited to nine as before. Third, we now estimate famine intensity at the oblast level using previously unavailable population reconstructions of famine related mortality. As before, we analyze T2DM outcomes by month of birth to establish critical time windows for early life famine exposure in relation to later health.

## METHODS

We used three variables to assess adult T2DM risk in relation to early famine exposure. First, the size of the population at risk as study denominator; second, the number of T2DM cases in later life as the study numerator; and third, excess mortality as an indicator of the level of famine intensity. These parameters were estimated for specific geographical units that show wide variations in the timing and intensity of famine exposure. We then examined potential dose-response relations between the level of prenatal famine in different periods of gestation and adult T2DM.

Ukraine is currently composed of 24 oblasts (provinces) and the Autonomous Republic of Crimea, annexed by Russia in 2014. Seventeen of these oblasts cover what used to be Soviet Ukraine and they show significant variation in the degree of famine exposure in 1932-1933. These oblasts were ordered in three groups (with significant, severe, or extreme famine) according to the level of famine exposure as determined by excess mortality during the famine.

The remaining seven oblasts cover what used to be Polish, Romanian, or Czech territory at the time of the famine and had no famine exposure. These oblasts were annexed by Soviet Ukraine after WWII. Crimea was omitted because it was not part of Soviet Ukraine during the Holodomor. Zaporizhzha oblast was omitted because of incomplete data.

### Population reconstruction to estimate excess deaths and actual births

Excess mortality is defined as the difference between the actual number of deaths during the famine period and the estimated number of deaths during the same period had there been no famine. The actual number of deaths during the Holodomor was much higher than the number of registered deaths. This requires an adjustment of registered deaths, especially in 1933.^3^ To assess estimated deaths had there been no famine, reliable estimates of mortality levels before and after the famine are needed. Elements for making these estimates are provided by the methodology of population reconstruction. This methodology was also used to estimate numbers of monthly births by oblast and sex from 1930 to 1938.

### Type 2 diabetes mellitus assessment

To ascertain T2DM outcomes among the populations at risk defined by sex, month and year of birth 1930-1938, and oblast of residence, we identified all T2DM cases included in the national Ukraine diabetes register (Komisarenko Institute of Endocrinology and Metabolism, Kyiv) for individuals born in Ukraine between 1930-1938. The register was created in 2000 and last updated in 2008. Reports by primary care physicians for diabetics are the primary data source for the register. T2DM diagnosis was based on WHO (1999) criteria.^12^ To minimize bias from Type One Juvenile Onset Diabetes contamination, we restricted cases to the 141,012 individuals in the registry diagnosed with T2DM at age 40 or over. For 12,787 individuals (9.1%), birth month was unknown, leaving 128,225 cases for further analysis.

### Famine intensity

Prior to data analysis, the 17 oblasts were grouped into three categories by their excess mortality: a) extreme famine (212 - 243 losses/1,000 population): Poltava, Cherkasy, and Kyiv; b) severe famine (97 - 161 losses/1,000): Vinnytsia, Zhytomyr, Kirovohrad, Sumy, Kharkiv, Kherson, Mykolaiv, Odesa, and Dnipropetrovsk; and c) significant famine (47 - 94 losses/1,000): Chernihiv, Khmelnytskyi, Luhansk, and Donetsk. The seven oblasts of Western Ukraine were not exposed to famine. The map shows great variations in the famine intensity across Ukraine, from 47 excess deaths per 1,000 population in Donetsk oblast to 243 excess deaths in Poltava oblast (**Figure 1**). The number of births in the first half year of 1934, nine months after the famine, in the regions with extreme, severe, significant, and no famine, was 22%, 32%, 46%, and 97% respectively compared to that during pre-famine periods. Therefore, famine-related changes in mortality were followed by proportional changes in fertility nine months later.

**Figure 1.**
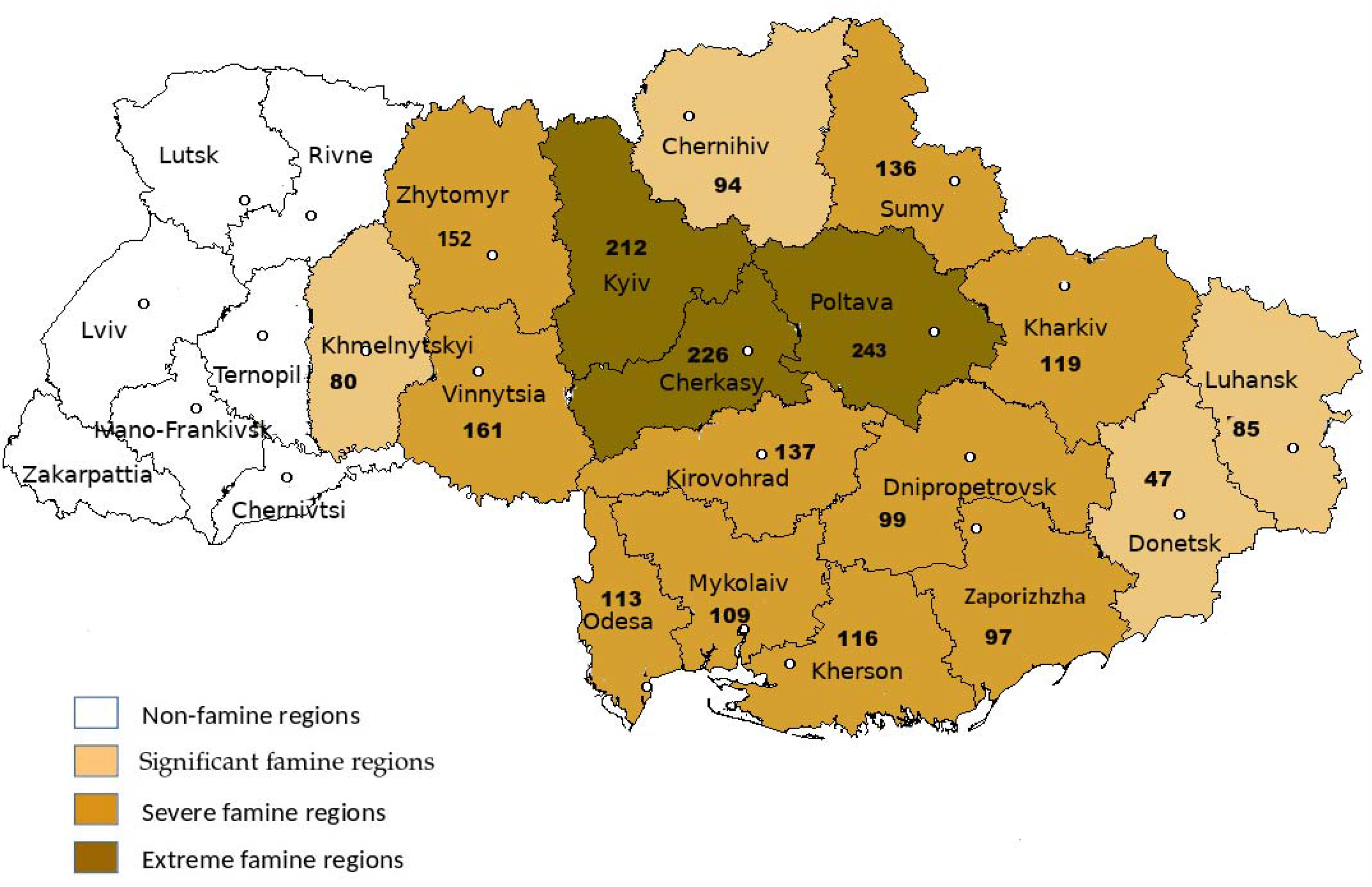
Regional distribution of 1932-1933 famine losses (per l,000 population) by level of famine: significant (47 to 94), severe (97 to 161) and extreme (212 to 243)

### Study conduct

The use of anonymized data was approved by the institutional review board of the Komisarenko Institute of Endocrinology and Metabolism, Kyiv, Ukraine, and the need for informed consent for the purpose of this study was waived.

### Statistical analysis

We estimated long-term famine effects using a Difference-in-Differences approach to capture interactions of oblasts and month of birth over time.^13,14^ As noted, the place classification identifies four region groups: oblasts with extreme, severe, significant or no famine losses. We examined differences in discrete outcomes (the odds of T2DM) on a multiplicative scale in logistic regression models. We carried out exploratory analysis as outlined in **Appendix** and identified a peak increase in T2D odds for births between January-June 1934 in famine exposed oblasts.

We then also combined all region groups to estimate the T2DM odds associated with births in extreme, severe, and significant famine regions as any famine exposure relative to births in no famine regions. We modeled T2DM as a function of region, year of birth, and season of birth and their interactions first for men and women separately and then combined, adjusting for sex.

In sensitivity analyses, we examined potential changes in the odds for T2DM using as controls only pre-famine births or only post-famine births or combining the two groups. We further conducted a meta-regression to integrate associations between famine intensity and T2D risk at the oblast level and the regional level.

## RESULTS

Our study population at risk for T2DM includes the 10,186,016 Soviet Ukraine births in the years 1930-38 outside of Zaporizhzha. 51.8% of births were men and 48.2% were women. 14.5% of the population was born in regions with extreme famine, 38.8% in regions with severe famine, 22.2% in regions with significant famine, and 24.4% in regions with no famine (**Table 1**). Study outcomes include the 128,225 T2DM patients from the national T2DM registry 2000-2008 who were born between 1930-38. Information on births and T2DM cases classified by sex, half-year and year of birth, and by region of famine intensity is provided in **Appendix Supplementary Table 1**.

**Table 1.** Odds of Type 2 diabetes mellitus (T2DM) 2000-2008 Reconstructed births in January-June 1934 vs all other Ukraine births in 1930-1938 by regional intensity of Ukrainian famine 1932-1933 Odds Ratios and 95% Confidence Intervals.

| **Diabetes Univariate analysis within birth region#** |  |  |  |  |  |  |
| --- | --- | --- | --- | --- | --- | --- |
| 128,225 T2DM cases (Men n=38,790 and Women n=89,435) |  |  |  |  |  |  |
| Births in 1st half year 1934 (January-June) vs all other births 1930-1938 |  |  |  |  |  |  |
|  | Men<br>n=5,272,731 |  | Women<br>n=4,913,285 |  | All<br>n=10,186,016 (adj. for sex) |  |
|  | OR | 95% CI | OR | 95% CI | OR | 95% CI |
| **Region of Birth** |  |  |  |  |  |  |
| Extreme Famine n=1,478,456 | 2.16 | 1.89-2.47 | 2.16 | 1.97-2.37 | 2.15 | 1.99-2.32 |
| Severe Famine n=3,955,120 | 1.98 | 1.83-2.14 | 1.92 | 1.82-2.03 | 1.93 | 1.85-2.02 |
| Significant Famine n=2,265,136 | 1.58 | 1.41-1.77 | 1.45 | 1.35-1.56 | 1.48 | 1.40-1.58 |
| No Famine n=2,487,304 | 0.94 | 0.85-1.04 | 0.92 | 0.86-0.99 | 0.93 | 0.87-0.98 |
| Any Famine n=7,698,712 | 1.85 | 1.75-1.96 | 1.78 | 1.71-1.85 | 1.80 | 1.74-1.85 |
| **Multivariable Analysis across birth regions\*** |  |  |  |  |  |  |
|  | Men<br>n=5,272,731 |  | Women<br>n=4,913,285 |  | All<br>n=10,186,016 (adj. for sex) |  |
|  | OR | 95% CI | OR | 95% CI | OR | 95% CI |
| **Region of Birth** |  |  |  |  |  |  |
| Extreme Famine n=1,478,456 | 2.45 | 1.93-3.11 | 2.37 | 2.00-2.80 | 2.39 | 2.09-2.74 |
| Severe Famine n=3,955,120 | 2.31 | 1.91-2.79 | 2.41 | 2.11-2.75 | 2.38 | 2.13-2.65 |
| Significant Famine n=2,265,136 | 1.92 | 1.54-2.39 | 1.95 | 1.68-2.26 | 1.94 | 1.71-2.19 |
| No Famine n=2,487,304 | Ref | - | Ref | - | Ref | - |
| Any Famine n=7,698,712 | 2.19 | 1.84-2.60 | 2.23 | 1.97-2.51 | 2.21 | 2.00-2.45 |

#Cross-product Odds Ratios within birth region
\*Difference in Differences analysis. Odds Ratio's for the 3-way interaction terms of the main effects year of birth (1934 vs all other years 1930-1938), half-year of birth (January-June vs July-December), and birth region (exposed to extreme/severe/significant famine) in a single model including all lower-level interaction terms and adjustment for sex where applicable.

### Exploratory analysis to identify critical time window

In exploratory analyses as described in **Appendix** T2DM prevalence showed the strongest increase among births in the first half-year of 1934. The increase was not seen among births in any of the months before or after. Specifically, there was no adult T2DM increase among individuals exposed to famine in the first year of life. Stratification by region of birth shows a dose-response relation, with the larger T2DM increase in regions exposed to more severe famine in 1933 (**Figure 2**).

**Figure 2.**
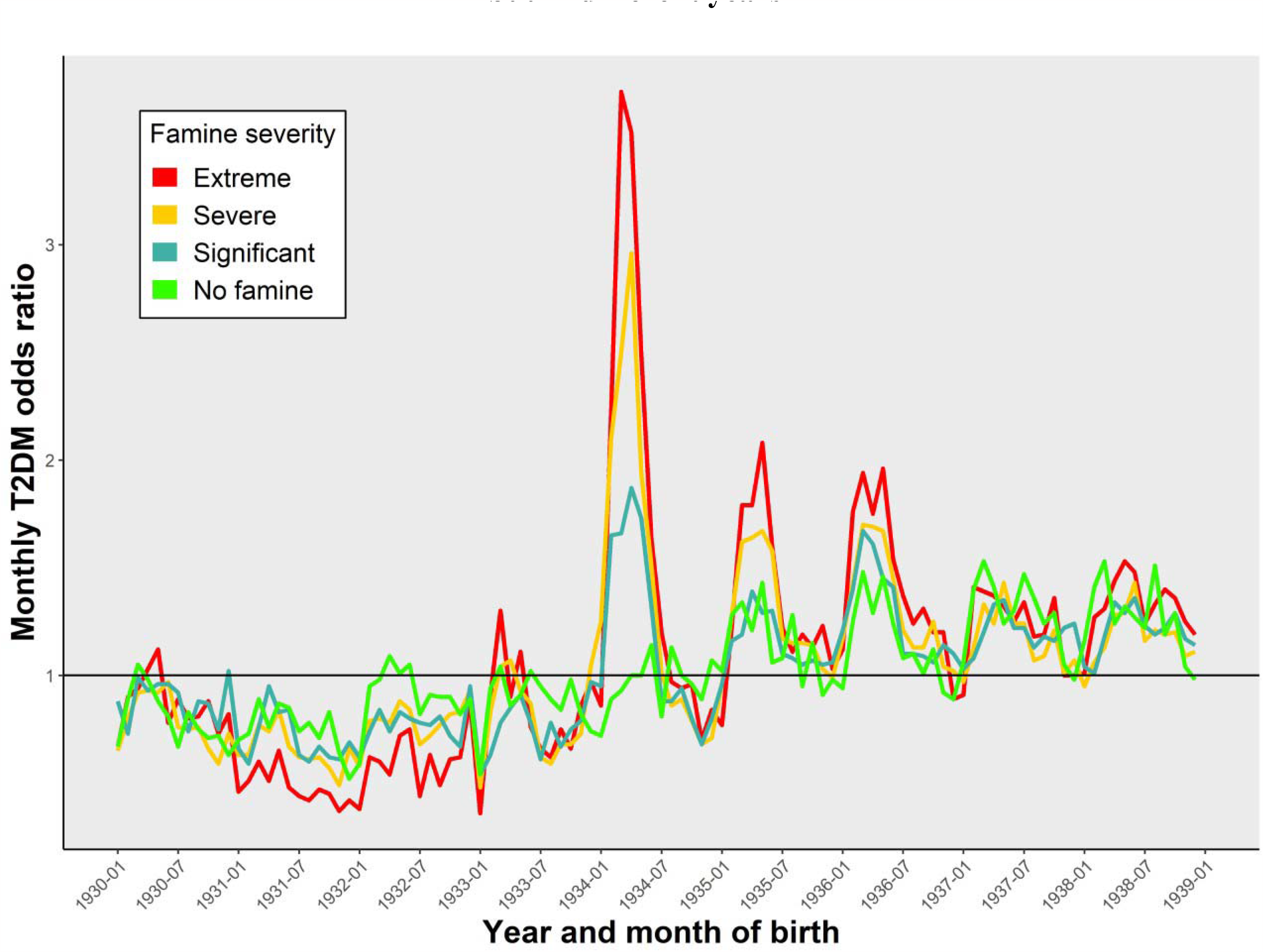
Odds of Type 2 diabetes mellitus (T2DM) among births in any specific month and year from January 1930 to December 1938 relative to births in the same month and region but in different years.

### T2DM increase in selected critical time window

Within regions of different famine intensity, births in January-June 1934 in regions with extreme famine showed a 2.15-fold increase in T2DM, in regions with severe famine a 1.93-fold increase, and in regions with significant famine a 1.48-fold increase relative to the pre-of post-famine births in the same region. Regions with no famine showed no T2DM increase compared to time-controls. Births after any famine exposure (combining extreme, severe, and significant regions) showed a 1.80-fold increase (95% CI: 1.74-1.85) in T2DM relative to unexposed controls. Findings in men and women were similar (**Table 1, upper panel**).

Comparing famine births to unexposed controls in multivariable analysis, births in regions with any famine exposure (extreme, severe, and significant famine combined) showed a 2.21-fold increase (95% CI: 2.00-2.45) in T2DM prevalence. Within the any famine exposure group, births in regions with extreme and severe famine showed a 2.39- and 2.38-fold increase respectively and births in regions with significant famine a 1.94-fold increase relative to births in regions with no famine. Again, men and women showed similar findings (**Table 1, lower panel**).

### Sensitivity analysis

Effect estimates did not differ using either pre-famine or post-famine births as controls and the groups were combined in final analysis. The relation between T2DM outcomes in each of the 17 oblasts and Western Ukraine in relation to excess mortality in 1933 is shown in **Figure 3**. The meta-regression shows a consistent trend at the oblast and at the regional level comparing famine intensity in early gestation and later T2DM.

**Figure 3.**
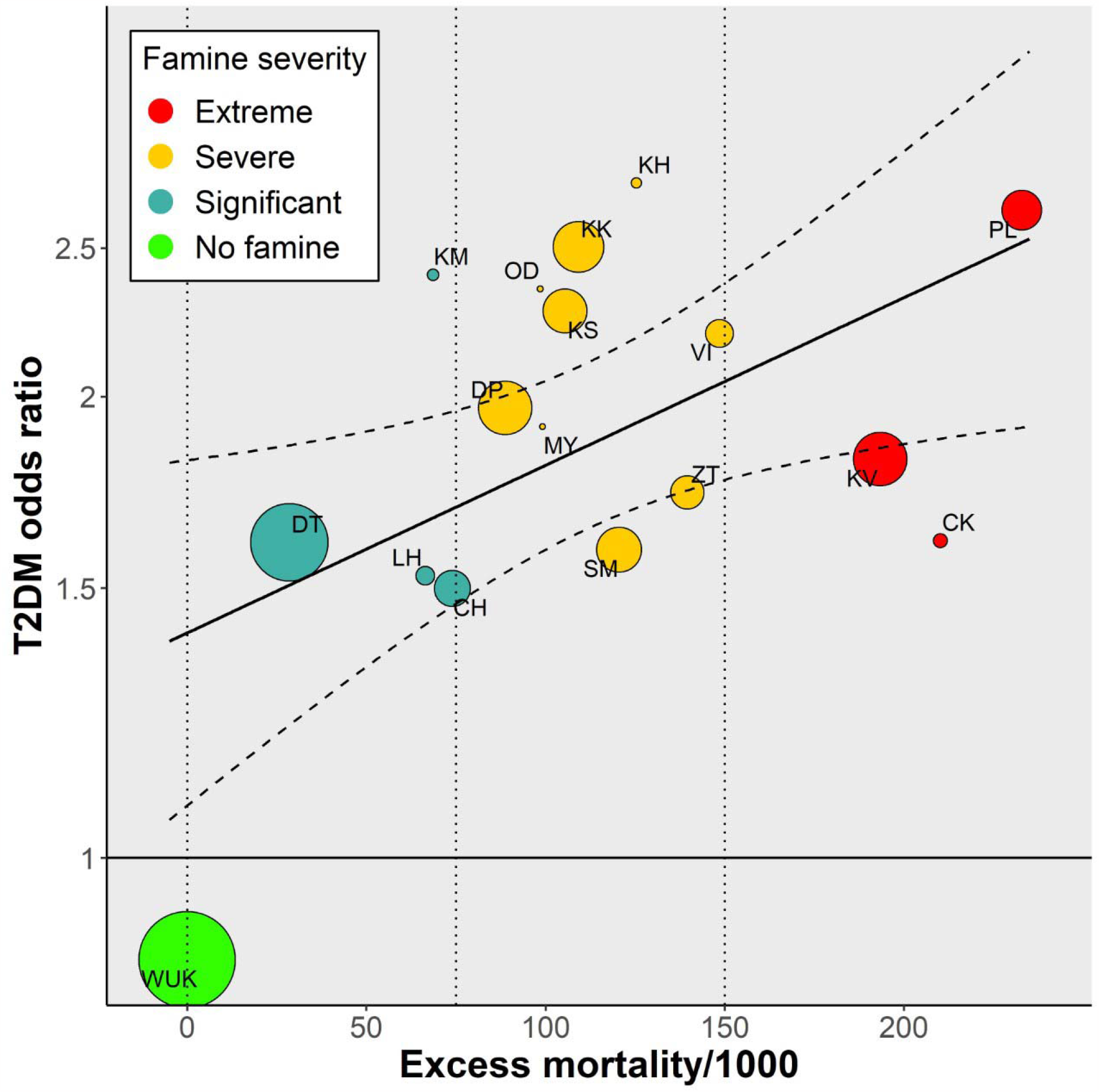
Association between famine intensity and T2D risk at the oblast level and with oblasts color coded by famine severity at the group level. The size of each dot is proportional to the weight of the study in terms of population size. The dashed colored lines represent the 95% Cl for the meta-regression model. Oblasts with extreme famine: CK:Cherkasy, PL:Poltava, and KV:Kyiv; with severe famine: DP:Dnipropetrovsk, KK:Kharkiv, KS:Kherson, KH:Kirovohrad, MY:Mykolaiv, OD:Odesa, SM:Sumy, VT:Vinnytsia and ZT:Zhytomyr; with significant famine: CH:Chernihiv, DT: Donetsk, KM:Khmelnytskyi, and LH:Luhansk; with no famine: WUK:Western Ukraine.

## DISCUSSION

Our findings show a larger than 2-fold increase in the odds of developing T2DM in later life in men and women born in famine exposed provinces in Ukraine in early 1934 (OR 2.21; 95% CI: 2.00-2.45). Stratification by level of famine severity shows a dose-response relation. These births were conceived in the 6-month period around June 1933 when famine mortality was at its highest. The T2DM increase in later life is therefore related to famine exposure in early gestation. No T2DM increase was seen among infants exposed to famine in mid or late gestation or in the first years of life. Outcomes in men and women were largely overlapping but are reported by sex and adjusted for sex for completeness.

The 2-fold T2DM increase is higher than the 1.5-fold increase reported in 2015 by some of us in a previous Ukraine study limited to nine selected oblasts.^9^ As an important limitation, the study population at the time was defined by individuals included in the 2001 national Census of Ukraine. The previous study was further limited to births in an opportunity sample of the nine Ukraine provinces for which data were then available. Our nationwide study including 23 out of the 24 oblasts avoids potential biases related to the selection of oblasts.

The well-defined temporal relationship between Holodomor births and adult T2DM emerges in part from the dynamics of the Ukraine famine. The Holodomor far exceeded other major famines in terms of its intensity, with an excess mortality reaching 140/1,000 overall and exceeding 200/1,000 in some oblasts. It resulted in 4 million excess deaths. In the Chinese famine of 1959-61, the largest famine of modern times, with an estimated 30 million excess deaths, national mortality did not exceed 25/1,000 but a much larger population was affected.^15^ The Holodomor losses were concentrated between January-June 1933 (for Mortality Surge see **Appendix)** while in China the famine losses were spread over several years. The Ukraine famine characteristics therefore allow for unusual specificity in defining the timing of exposure.

The assembly of Ukraine reconstructed births between 1930-38 as study denominator for the population at T2DM risk overcomes limitations associated with the previous use of the 2001 Census population for this purpose. The reconstructed births include 10.2m individuals of which 52% were male. The 2001 Census population for births between 1930-38 includes 3.6m individuals of which 40% were male. This indicates that 27% of men and 43% of women of the birth cohorts were no longer represented in the census, either because they were no longer alive or because of other reasons. These losses could give rise to multiple biases of unknown magnitude and direction, including from premature deaths related to T2DM or other conditions. Our observations on cohort survival are in agreement with the 27% male and 49% female survival rates seen in Ukrainian cohort life tables for 1932-1933 births.^16^

With regard to classifying famine severity experienced in early life, the meta-regression of famine intensity and later life T2DM based on oblast level findings (**Figure 3**) confirms the suitability of our pre-study arrangement of oblasts in four grouped categories (extreme, severe, significant, or no famine exposure) (**Table 1**). While there is variation in the relation between oblast famine intensity and later T2DM in individuals, the overall picture shows a robust association between famine intensity and later life T2DM. We note that the T2DM association in oblasts exposed to extreme or severe famine shows considerable overlap and that these categories could be merged without loss of information. As discussed below, we have no indications that measurement errors of famine intensity or diabetes prevalence differed by oblast.

### Strengths and limitations

Among the strengths of our study we mention the successful assembly of all pre-famine, famine, and post-famine monthly births between the years 1930-38 in Ukraine oblasts to define populations at risk; the famine exposure defined by place by the multi-level classification of famine severity at the oblast and regional level; the unexposed place controls provided by Western Ukraine oblasts that were not part of Soviet Ukraine during the famine; the famine exposure defined in time by specific characteristics of this man-made famine that resulted in extreme human losses condensed in a 6-month time period; and the famine outcomes ascertained nationwide through the 2000-08 Ukraine national diabetes register after a long follow-up period.

We also note some limitations. First, famine exposure was estimated at the province level as measures of famine exposure at the individual level are not available. Second, T2DM cases in the national register were reported by oblast of current residence. And third, the diabetes register cannot be used to reliably determine T2DM prevalence in Ukraine because cases are underreported. We discuss the potential impact of these limitations in **Appendix**.

More generally, the study cannot exclude potential confounding by selected lifestyle characteristics or behaviors. As an example, we have no information on income, education, or dietary habits that could drive T2D risk and act as potential mediators of T2D risk. And information on being overweight or obese which could be an important factor in determining diabetes is not available.^6,17^ Any of the above if significantly different in births between January and July 1934 in famine affected areas could be a potential threat to the study’s validity.

### In search of mechanisms

To date, studies in humans have only been able to provide a general direction regarding possible mechanisms related to early gestation famine exposure. In temporal order these are reflected in selective survival of fetuses,^18^ DNA methylation changes at the imprinted IgF2 gene^19^ or candidate genes involved in metabolic disease,^5,20-22^ increased overweight in young adults, increased T2DM in middle age, and increased mortality through age 65.^6,13,23^ Further exploration will likely require experimental studies in animals to examine specific hypotheses.

### Implications of findings, also for current situation in Ukraine

The current Russian war against Ukraine has generated both immediate and far-reaching implications on global food security, particularly affecting some countries in Africa and Middle East which heavily depend on food imports from Ukraine and Russia.^24^ While the issue of food scarcity was already a persistent concern in these countries, the Russian invasion of Ukraine has substantially exacerbated this critical challenge and could lead to an increase in famine-related deaths and long-term adverse health outcomes.

## Data Availability

All data produced in the present work are contained in the manuscript.

## ACKNOWLEDGEMENTS

Funding provided by the Ukraine State complex program Diabetes Mellitus, project number 0106U000844. LHL and OW received support from the Holodomor Research and Education Consortium in Canada. LHL received support from a NIDI-NIAS Fellowship of the Royal Netherlands Academy of Sciences and from grants R01 AG028593 (PI: Lumey) and R01 AG06687 (MPIs: Lumey and Belsky) National Institute of Aging, National Institutes of Health. We thank Dr. Vitaliy Gurianov (Bohomolets National Medical University, Kyiv, Ukraine) for statistical advice.

