## Supplementary material for "Long-term impact of pre-natal exposure to the Ukraine famine of 1932-1933 on adult type 2 diabetes mellitus": Ukraine Lumey et al. Supplementary materials.docx

**APPENDIX**

**Population Reconstruction: Overview**

**Use of the demographic equation**

Excess mortality is defined as the difference between the actual number of deaths during the famine period and the estimated number of deaths during the same period had there been no famine. The actual number of deaths during the Holodomor was much higher than the number of registered deaths. This requires an adjustment of registered deaths, especially in 1933.^1^ Estimating expected deaths had there been no famine requires reliable estimates of mortality levels before and after the famine. Elements for making both of these estimates are provided by the methodology of population reconstruction based on the demographic equation:

*P( t* + 1) = *P(* t) + *B( t, t* + 1) - *D( t, t* + 1) + *l\M( t, t* + 1), where

*P* = January 1^st^ population of year *t*; *B* and *D* = number of births (and deaths) during year *t* and *NM* = number of in-out-migrants during year *t*.

Starting with the initial census population, successive yearly populations are obtained by adding the number of births, subtracting the number of deaths, and adding the number of net migrants for each year until the post-famine census year is reached.

The equation provides the number of actual deaths during the famine period. The estimation of the expected number of deaths during the famine period had there been no famine is done in two steps: a) interpolating death rates before and after the famine; b) multiplying the interpolated death rates for the famine years by the respective (sub)populations.

### Birth cohorts at risk

Population reconstructions also generate estimated numbers of yearly births by oblast and sex from 1930 to 1938. As for excess mortality, reconstructed births were originally calculated for the seven oblasts of Soviet Ukraine, then recalculated for the current 17 oblasts using the same transition coefficients as for excess deaths. Yearly births for oblasts of Western Ukraine were estimated using vital statistics and census data from three different national statistic systems: Poland, Romania, and Czechoslovakia. The next step was to decompose the yearly births by month. The result is a complete series of monthly births by sex for 23 of the 24 oblasts of Ukraine, from 1930 to 1938.

### Famine intensity

Famine losses were originally estimated in the period 1932-34 for the seven oblasts of Soviet Ukraine at the time of the Holodomor, with a total of 3.9 million excess deaths.^2^ Famine losses were recalculated for the current 17 oblasts that encompass the territory of former Soviet Ukraine, using transition coefficients based on the population of respective fractions of oblasts and expressed as the number of excess deaths per 1,000 population.

**Population Reconstruction: Details**

**Estimation of yearly deaths and births for Soviet Ukraine.**

Yearly reconstructed populations were estimated for the 17 oblasts of Soviet Ukraine for the period January 1, 1927 to January 1, 1938. Data for the 1930-38 period containing the Holodomor years were used in our analysis. The initial and final populations of this period are based on the 1926 and 1939 censuses. Data from the 1937 census and the 1931 urban count were used as controls in the population reconstruction process. This methodology is data intensive and requires yearly time series of births, deaths and net migration for the reconstruction period.

A key element of the population reconstruction methodology is an initial evaluation of each component of the demographic equation and making relevant adjustments where necessary. For example, the 1926 and 1937 censuses required minor adjustments, but the 1939 census required major adjustments because it was falsified to minimize the population losses caused by the Holodomor.^3^ As a result, our adjustments reduced the official total population of Ukraine by 2.6 percent.^1^

Registered births and deaths required minor adjustments during the non-famine years, but many deaths and births were not registered during the Holodomor, especially in 1933. For example, it has been estimated that about one-third of births and more than half of deaths were not registered in 1933.^1^ In addition, net migration had to be estimated piecemeal, as no comprehensive information on migration was available. These adjustments were refined further using the demographic equation and adjusted census populations as controls.

**Estimation of births for non-famine regions**

1932, 1933 and 1934 births for the seven oblasts of Western Ukraine were estimated using vital statistics and census data from three countries. We used transition coefficients derived by comparing relevant maps and census populations at the regional levels of these countries - povity in Poland and Romania and okruhy in Czechoslovakia - with maps of the seven oblasts. Total yearly births were then adjusted for under-registration using methods described.^1^ Yearly total births for the non-famine years, 1930-31 and 1935-38, were taken from the population reconstruction of Western Ukraine done by O. Rudnytskyi for the 1795-1959 period.^4^

**Estimation of Holodomor losses and births by month**

*Soviet Ukraine*

Registered births by month for the seven original oblasts are available only for 1932, 1933 and 1934 during the 1930-38 period. The number of births by month for the other years is only available for the whole country. We estimated the number of births for the 17 oblasts by month for the non-famine years by applying the respective monthly distribution of births for the whole country to all oblasts.

*Western Ukraine*

Births by month for the oblasts of Western Ukraine are available only for 1932, 1933, and 1934. Therefore, the monthly distributions of births were calculated separately for: a) the five western oblasts that were part of Poland at that time; b) Chernivtsi oblast, which was part of Romania; c) Transcarpathian (Zakarpattia) oblast, which was part of the Czechoslovak Republic.

Two separate estimation methods were applied to the five oblasts that were part of Poland. The monthly proportions of births of the group of southern voivodships of Poland were used to estimate births by month for Ivano-Frankivsk, Lviv, and Ternopil oblasts. Monthly births for Volyn and Rivne oblasts were estimated using the monthly structure of births of the eastern voivodships of Poland. Yearly births in Chernivtsi oblast were distributed by month using monthly birth statistics from the respective counties of Bessarabia and Bukovyna (regions of Romania at that time). Monthly births in Transcarpathia were estimated by applying the monthly distribution of births in Carpathian Ruthenia (part of Czechoslovakia at that time). Due to a lack of data, the monthly distribution of births for the non-famine years were estimated by dividing the total number of yearly births by 12.

**Mortality surge in early 1933**

A unique characteristic of the Holodomor is that most losses are concentrated in a short period. Three and a half million, or 89.5 percent of the 3.9 million 1932-34 Holodomor losses happened in 1933. Of the 3.5 million losses in 1933, 84 percent occurred during the year's first seven months (Figure).^5^

Holodomor losses experienced an extraordinary increase between January and June 1933. The number of excess deaths in Soviet Ukraine increased almost ten times between January and June, from 87,718 to 840,703. At the Famine's peak in June 1933, there were, on average, 28,000 Holodomor-related deaths per day.

**Figure. Percent Monthly Famine Losses for 1932-34**

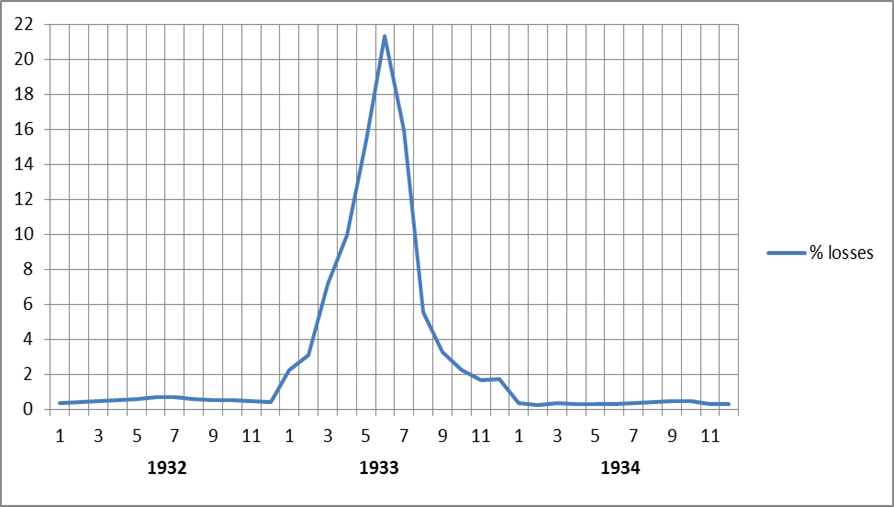

Source: author’s calculations.

**Exploratory Statistical Analysis**

In exploratory analyses, we first examined in each region group if the odds for T2DM were elevated in any of the birth months in the period January 1930 to December 1938. This was done by comparing the region specific T2DM odds for each year and month of birth relative to the same months of birth in all other years combined. This stratified analysis serves to identify potential year of birth effects controlling for month of birth effects. We observed a peak increase in T2D odds for births between January-June 1934 in famine exposed oblasts and smaller increases for births in 1935 and 1936 in the same months. This demonstrated that statistical control for month of birth effects could be achieved by adjusting for the January-June period in multivariate modelling.

**Study Limitations**

**Famine exposure was estimated at the province level** **as measures of famine exposure at the individual level are not available.** It is not possible therefore to determine what minimum nutrition level in pregnancy is essential to avoid long-term health effects for the newborn. Average individual and group-level exposures will be similar however if well-defined famine areas are separated from non-famine areas by natural or man-made borders and leaving the famine area is not possible. This was the case in Ukraine when fleeing famine areas was hindered because of the closing of borders by Soviet officials. Major factors in the mortality surge in early 1933 in Ukraine were the confiscation of all foods in rural households during searches of ‘hidden’ grain and targeted measures that made it almost impossible for peasants to travel in search of food.

**T2DM cases in the national register were reported by oblast of current residence**. We used this as a proxy for oblast of birth. While relocations within oblast of birth will not change study results, out-migrations if not counterbalanced by return migrations might. To evaluate the magnitude of this potential problem, we compared province of birth 1930-38 to province of residence as both were collected in the 2001 Census. There had been no change in over 80% of the population. The agreement in Kyiv oblast was as high.^6^ In view of this high concordance, we used oblast of residence in 2001 as a proxy for place of birth 1930-38. Random misclassification of province of birth in the remaining 20% of the population could have biased the study results downwards.

**The diabetes register cannot be used to reliably determine T2DM prevalence in Ukraine.** Most patients with type 2 diabetes do not immediately have clinical symptoms that lead to detection and reporting. Between 2000-08, about 600,000 T2DM cases were entered into the registry, and this represents about half the number of cases that would be expected from Health Ministry Surveys and other sources. Registration of insulin treated cases is the most complete (~90%) as insulin treatment for registered T2D patients is provided free-of-charge in Ukraine.

The register is highly suitable however to examine famine related T2DM differentials between oblasts if the reporting coverage is comparable across oblasts. T2DM prevalence data from the Ukraine Ministry of Health in 2014 (not broken down by subtype or treatment modality) for the 27 current Ukraine oblasts show that oblasts reported similar T2DM prevalence, with variations no larger than 25% from the national mean.^7^ This justifies the use of the diabetes register as an indicator of between oblast T2DM prevalence.

**Supplementary Table 1. Ukraine births and registered T2DM cases 2000-08**

**by sex, famine intensity in region of birth, and half-year of birth 1930-38**

**(excluding Zaporizhzha)**

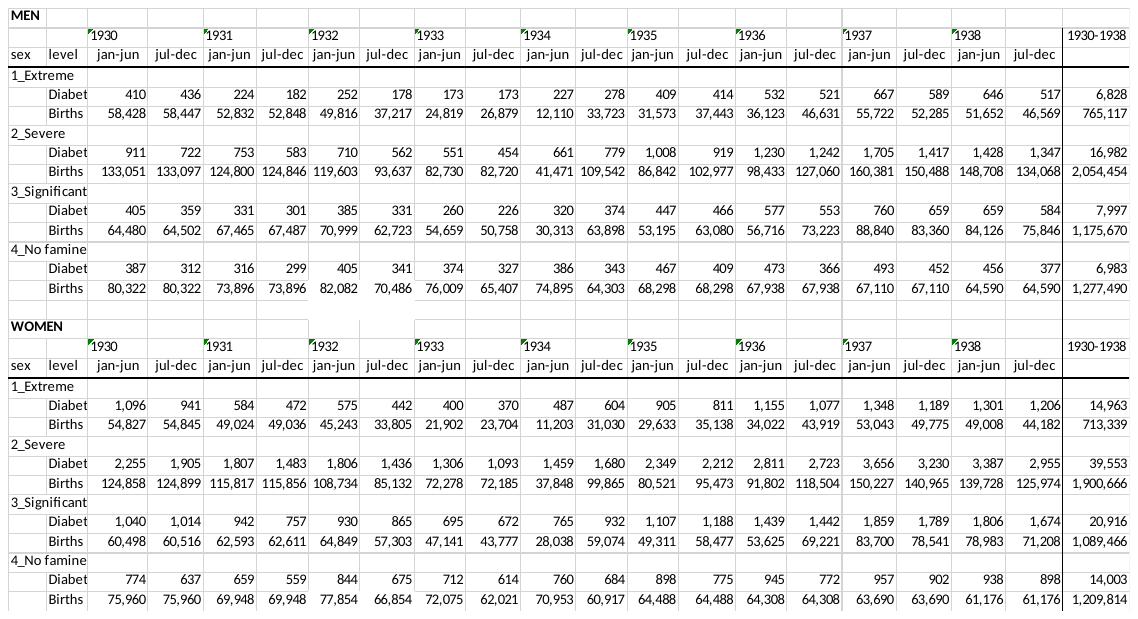
